# Can process mapping and a multi-site Delphi of perioperative professionals inform our understanding of system-wide factors that may impact operative risk?

**DOI:** 10.1101/2022.04.21.22274014

**Authors:** DJ Stubbs, T Bashford, FJ Gilder, B Nourallah, A Ercole, N Levy, PJ Clarkson

## Abstract

**Objectives:** To examine whether the use of process mapping and a multidisciplinary Delphi can identify potential contributors to perioperative risk. We hypothesised that this approach may identify factors not represented in common perioperative risk tools and give insights of use to future research in this area.

**Design:** Multidisciplinary modified Delphi study

**Setting:** Two centres (one tertiary, one secondary) in the United Kingdom during 2020 amidst coronavirus pressures.

**Participants:** 91 stakeholders from 23 professional groups involved in the perioperative care of older patients. Key stakeholder groups were identified through the use of process mapping of local perioperative care pathways.

**Results:** Response rate ranged from 51% in round one to 19% in round three. After round one, free text suggestions from the panel were combined with variables identified from perioperative risk scores. This yielded a total of 410 variables that were voted on in subsequent rounds. Including new suggestions from round two, 468/519 (90%) of the statements presented to the panel reached a consensus decision by the end of round three. Identified risk factors included patient level factors (such as ethnicity and socio-economic status); and organisational or process factors related to the individual hospital (such as policies, staffing, and organisational culture). 66/160 (41%) of the new suggestions did not feature in systematic reviews of perioperative risk scores or key process indicators. No factor categorised as ‘organisational’ is currently present in any perioperative risk score.

**Conclusions:** Through process mapping and a modified Delphi we gained insights into additional factors that may contribute to perioperative risk. Many were absent from currently used risk stratification scores. These results enable an appreciation of the contextual limitations of currently used risk tools and could support future research into the generation of more holistic datasets for the development of perioperative risk assessment tools.

**Strengths and Weaknesses:**

- Novel use of process mapping to identify key perioperative stakeholders
- Multidisciplinary Delphi panel to gain breadth of perspective
- Performed across two sites
- Comprehensive results may be of use to other researchers designing perioperative research databases
- Results may be limited by low response rate in final round (although majority of consensus decisions made in round two)

## Introduction

Understanding, predicting, and communicating risk is fundamental to perioperative practice [1]. The use of surgical risk stratification tools is a cornerstone of programs such as the National Emergency Laparotomy Audit (NELA) [2]. When using the output of such tools in day-to-day practice it is important to remember that they are developed from external electronic datasets and their application to decisions at the level of an individual centre or patient is complex and fraught with difficulty [3]. Risk scores commonly consist of patient and surgical factors that are statistically associated with relevant outcomes [4]. However, performance of such scores is far from perfect [5]. This raises the question as to whether currently unmeasured factors may improve our understanding and prognostication of risk. There is strong evidence that this is the case. Unwarranted variation is widespread within the National Health Service (NHS), with between-centre differences leading to discrepancies in cost, efficiency, and most importantly, patient outcome [6]. Significantly, organisational or system factors have been shown in a recent analysis of NELA data to be associated with worse outcomes [7]. However, such factors do not feature in commonly used risk assessment tools

Healthcare is increasingly digitised with electronic health records (EHRs) increasingly capturing detailed events from across the hospital system. In the perioperative setting, EHRs may hold data pertaining to an individual’s baseline state, operation details, physiological responses under anaesthesia, and perioperative complications. EHRs therefore appear to offer an appealing substrate to identify and test factors associated with perioperative outcome. However, in reality, due to the complexity of modern healthcare, the data they hold doesn’t accurately capture the ‘true’ patient state but the record is in fact biased by the care processes involved in the recording of such data [8]. To use EHRs to identify nascent factors that may broaden our understanding of perioperative risk, including at the level of the healthcare system, it is vital that we understand how that care is delivered, and the affect this might have on the electronic record. However, electronic records are themselves complex, with a multitude of both implicit and explicit data types.

A system may be defined as a collection of elements serving a common purpose, with emergent behaviour arising through their interaction. Such systems may be considered as simple, complicated, or complex, with complex systems exhibiting certain key behaviours such non-linearity, non-scalability, path dependence and emergence[9]. Healthcare has been posited as just such a complex system, with a perioperative system possibly including pre-assessment clinics, operating theatres, staff from multiple disciplines, patients, equipment, consumables, care processes, and culture. All of these may generate data which could provide novel insights into our understanding of risk. Researchers in other healthcare settings, where similar causal pathways may exist, have already recognised that standard statistical techniques may not be comprehensive enough to interrogate and understand such systems [10]. Therefore there is a need to consider additional strategies such as those used to understand and design other complex systems [11,12].

Given the complexity of healthcare, our aim in this paper was to employ a systems approach [11] to develop a holistic dataset that a breadth of perioperative professionals felt captured all dimensions of perioperative risk. The output of such an endeavour would enable a rational approach to extracting information from EHRs for future research and enable a clearer understanding of how this data is captured as part of the wider care process. Such strategies as well as offering a clearer scientific rationale for future hypothesis testing, could also encourage better data governance – by only extracting data fields from the EHR that were felt to be of clear importance.

To develop our consensus dataset, we employed a stepwise approach. Firstly, we sought to visualise how perioperative care is delivered through process mapping. This is a technique which enables complex systems to be visualised as a series of steps representing decision-making points and pathways that has been used in various healthcare settings [13,14]. These process maps were then used to identify the range of perioperative professionals involved in patient care, and whose views we needed to capture. Using this list of ‘stakeholders’ we next conducted a modified Delphi [15] across two hospital sites seeking to gain consensus on the breadth of factors at both patient, operation, and system level that were felt to impact on patient outcome.

## Methods

### Setting and approvals

Participants were recruited from two UK hospital trusts utilising EHRs. Cambridge University Hospitals Trust (CUH) is a tertiary referral centre offering secondary and tertiary level surgical services to patients across the East of England. The West Suffolk Hospital (WSH) is a district general hospital based in Bury St Edmunds. WSH offers a range of secondary care services whilst referring patients for tertiary care to specialist centres including CUH. This study forms part of the ‘Designing Improved Surgical Care for Older people’ (DISCO) study, approved by the London and Surrey Borders Research Ethics Committee (reference 19/LO/1648). DISCO is jointly sponsored by the University of Cambridge and CUH.

### Methodological approach and techniques

Systems engineers use a variety of techniques to interrogate and design complex systems. A framework for applying these tools to the healthcare setting has been recently jointly published by the UK’s Royal Academy of Engineers, Royal College of Physicians, and Academy of Medical Sciences [11]. In this study we utilised brainstorming interviews and graphical elicitation[16], process mapping, and a Delphi process to ensure we captured the views of professionals (stakeholders) from across the perioperative system in two distinct sites. A project flow diagram demonstrating the sequential use of these techniques is shown in **Figure 1**.

**Figure 1:**
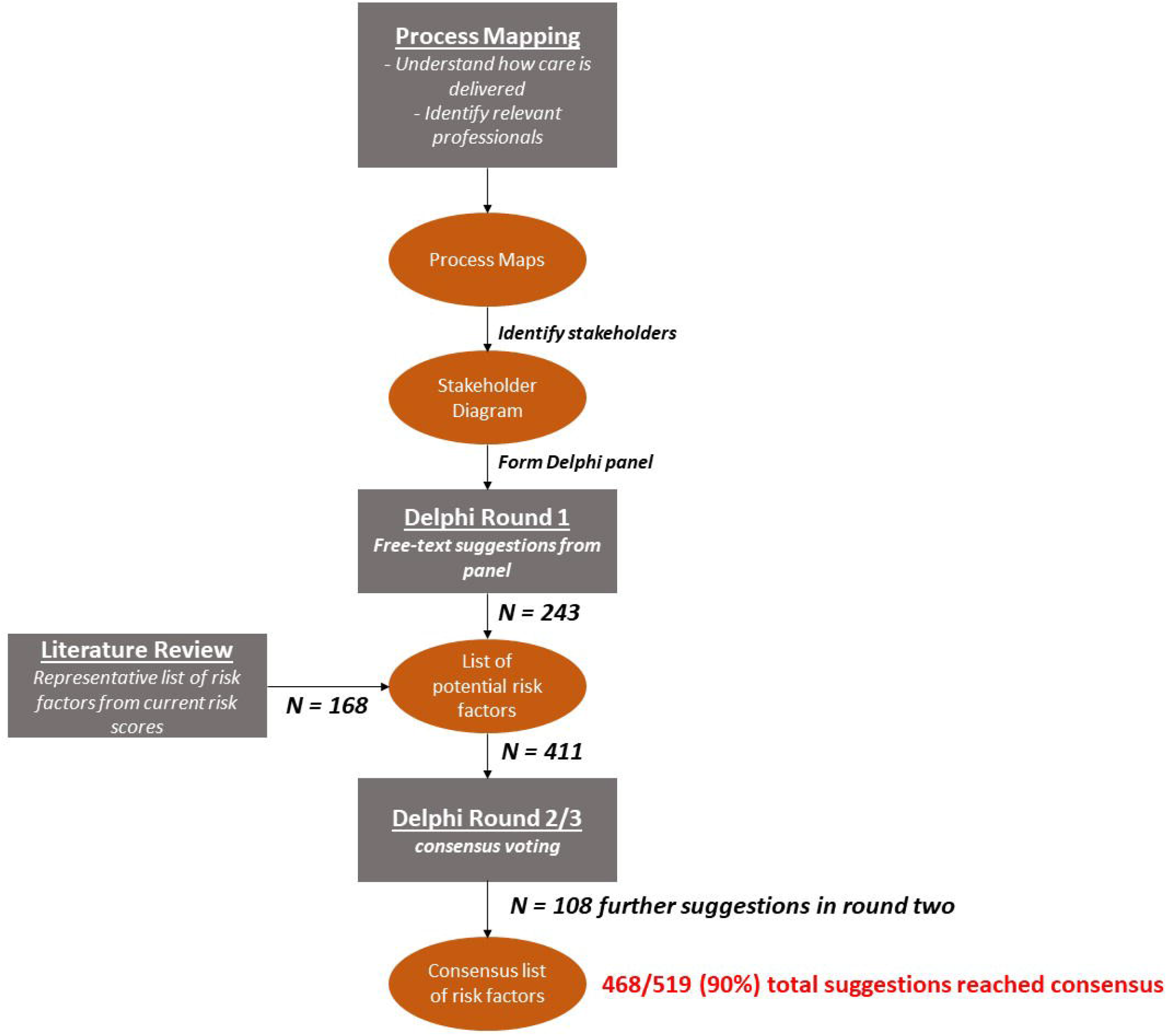
Project stages. Process mapping was performed with a steering group of experienced professionals, with maps iterated between brainstorming interviews. At the conclusion of this stage two outputs were generated – the maps themselves and a stakeholder diagram showing the viewpoints we sought to capture in our Delphi. The Delphi was conducted across three rounds with the first seeking free-text suggestions from the panel. These were combined with risk factors featuring in a systematic review of common perioperative risk tools [4]. All suggestions were then voted on by the panel across two further rounds to gain consensus. *N=* number of risk factors gained from each source. Our final consensus list of risk factors is available in **supplementary material**.

### Stakeholder identification and process mapping

We formed a local steering group of experienced perioperative professionals. The group consisted of a consultant anaesthetist, geriatrician, and surgeon alongside a senior physiotherapist, occupational therapist, matron, and operations manager. Brainstorming interviews were conducted to identify stakeholder groups who should be represented on the Delphi panel. Interviews were structured around the iteration of process maps representing a stereotypical ‘high risk’ surgical patient undergoing vascular surgery. Vascular surgery was chosen due to the ability to draw comparisons between elective and emergency cases as well as the need for clinical input from a range of perioperative professionals. Stakeholders were identified from these maps and then chosen for representation on the Delphi panel if at least one member of the steering group felt this was appropriate.

### Delphi panel formation

Representatives were approached from each stakeholder group across both sites aiming for a complementary spread of sub-specialities between sites. Individuals were approached by lead researchers in each site, provided with written information, and gave informed consent prior to each Delphi round.

### Delphi structure – round one

The Delphi consisted of three rounds and was distributed using an online survey tool (Qualtrics - www.qualtrics.com). In round one individuals were asked to provide free-text suggestions on what they felt *‘Could contribute to a poor-outcome in an older patient undergoing surgery’*. We defined a poor-outcome to be where an individual lost their independence after surgery or suffered a complication (such as a myocardial infarction). Suggestions from the panel were combined with known important risk factors identified from a systematic review of perioperative risk scores [4]. All risk factors (literature and panel suggestions) were voted on in the second and third rounds using a five-point Likert scale. Participants could also provide free-text comments and clarifications. A minority of questions in round two asked for specific free-text responses to expand on suggestions or provide relevant cut-off points (e.g. frailty score thresholds). New suggestions from round two were included for voting in round three. Before each major group of suggestions (e.g. ‘comorbidities’) respondents were asked to indicate on the same five point Likert scale how ‘measurable’ suggestions within this category were felt to be. This was to gauge the feelings of the panel on the practicality of this information being captured within an electronic record..

### Delphi structure – rounds two and three

All potential risk factors were presented for voting in a hierarchy consisting of groups and subgroups reflecting free-text suggestions. There were three broad domains - patient level factors (*that may have existed prior to an individual’s current admission: e*.*g. comorbidities)*, admission level factors (*the circumstances and events occurring in any given admission*), and system level factors (*suggestions that were related to the structure or running of a service within a healthcare organisation*). Tracking of free text data for generating round two and three questionnaires was conducted using ATLAS.ti (www.atlasti.com) with quantitative analysis conducted in R v 3.6.3 [17].

### Definition of consensus

Consensus was defined using criteria modified from a Delphi of quality indicators for patients with traumatic brain injury [50]. Given the relative heterogeneity of our panel, consensus for a given question required at least 50% of respondents to address it, a median score of >3.5, and an interquartile range (IQR) of ≤ 1. Consensus exclusion at the end of round two was stricter, with this requiring a score of <2.5, an IQR of ≤ 1, and no scores of 5 (‘very important’).

### Comparing to common literature sources

At the conclusion of the Delphi all novel, consensus, suggestions were compared to a systematic review of perioperative structure and process indicators[18] as well as whether they had been examined in the statistical development of each risk score[19–32]. This approach was chosen to ensure that our final list of factors contained a solid core of important patient level factors whilst allowing us to critique novel findings against a comprehensive literature source.

### Groupings, definitions, and results reporting

Certain questions in the Delphi allowed participants to vote on cut-points (e.g. “*a ‘high-risk’ body mass index (BMI) is above 30*”) or on an overarching concept that could encompass multiple factors (for instance ‘complications’ could encompass ‘perioperative myocardial infarction’ as well as surgical complications). For transparency all questions are presented in the **supplementary spreadsheet file** and included in relevant numerators or denominators within the results section. However, these cut-offs were not considered when reporting on factors present in literature risk scores unless they were explicitly mentioned (e.g. a risk score defined a specific cut-off).

### Patient and public involvement in project

The protocol for our project was reviewed by the CUH patient and public involvement panel in November 2017 and received favourable responses. The aims of our project are clearly aligned with the James Lind associations priority setting partnership conducted with the National Institute of Academic Anaesthesia [33].

## Results

### System mapping

Process maps representing the care of an elective and emergency vascular surgical patient were generated based on an understanding of local care pathways (abridged version in **Figure 2**, full examples as **supplementary figures 1 & 2**). These diagrams were revised based on brainstorming interviews with our steering group. Maps were used to identify relevant professional roles (stakeholders) across the perioperative pathway. In total, 52 unique staff stakeholder groups were identified. Of these, 33 were nominated by at least one member of the steering group for representation on the Delphi panel (**Figure 3**).

**Figure 2:**
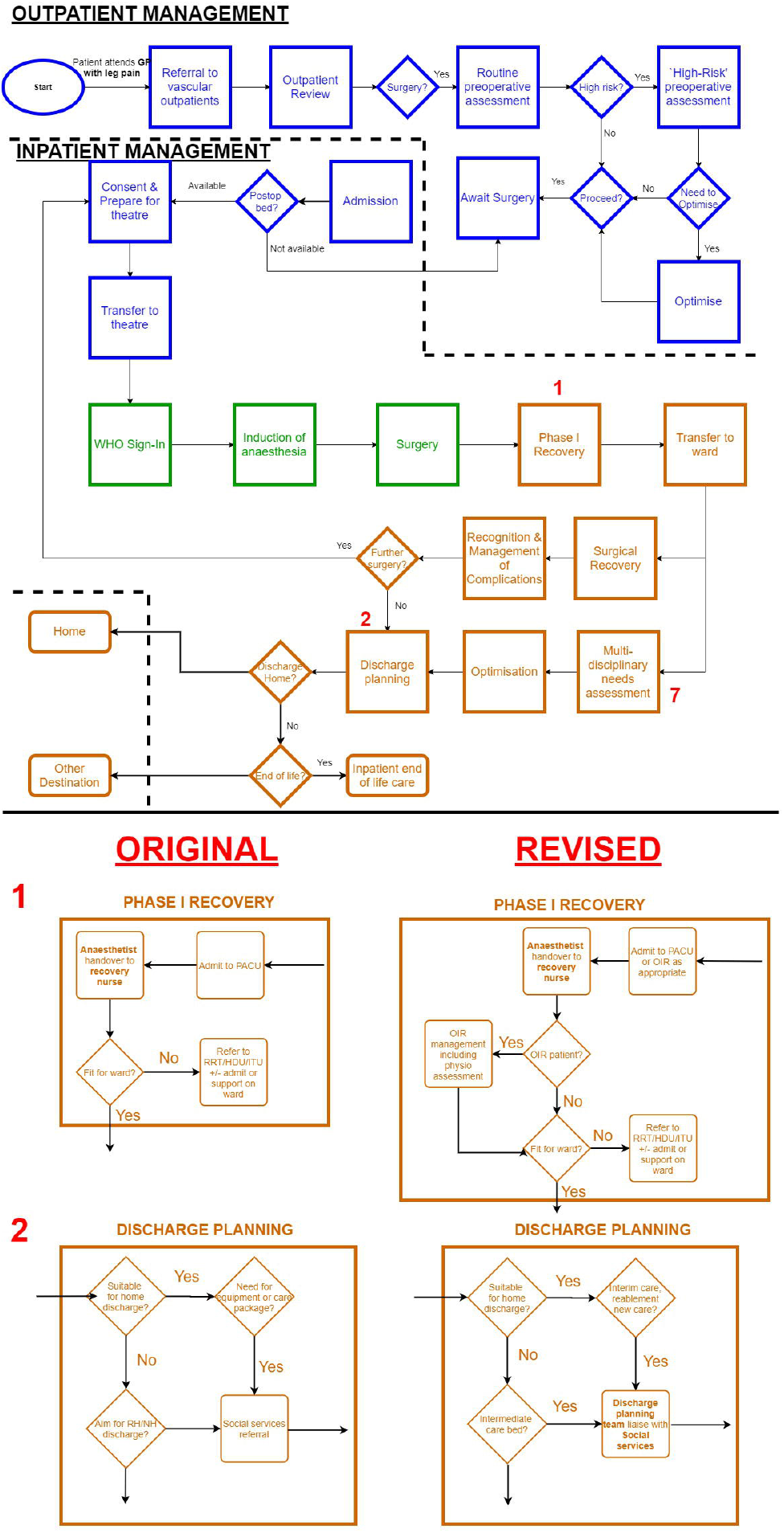
Simplified process map of a patient referred for elective vascular surgery at our institution. *Blue* boxes highlight preoperative care, *green* intraoperative care, *orange* postoperative care. Boxes indicate processes, diamonds indicate decision points. Dashed lines indicate boundary between community and hospital. Stakeholders (e.g. Anaesthetist) are shown in bold text. Lower panel highlights more granular view of numbered processes including revisions to initial diagram following discussions with a steering group of perioperative professionals. Fully granular maps for elective and emergency pathways are available in the supplementary material. *OIR = overnight intensive recovery (level 2 post anaesthesia care unit for high risk patients)*.

**Figure 3:**
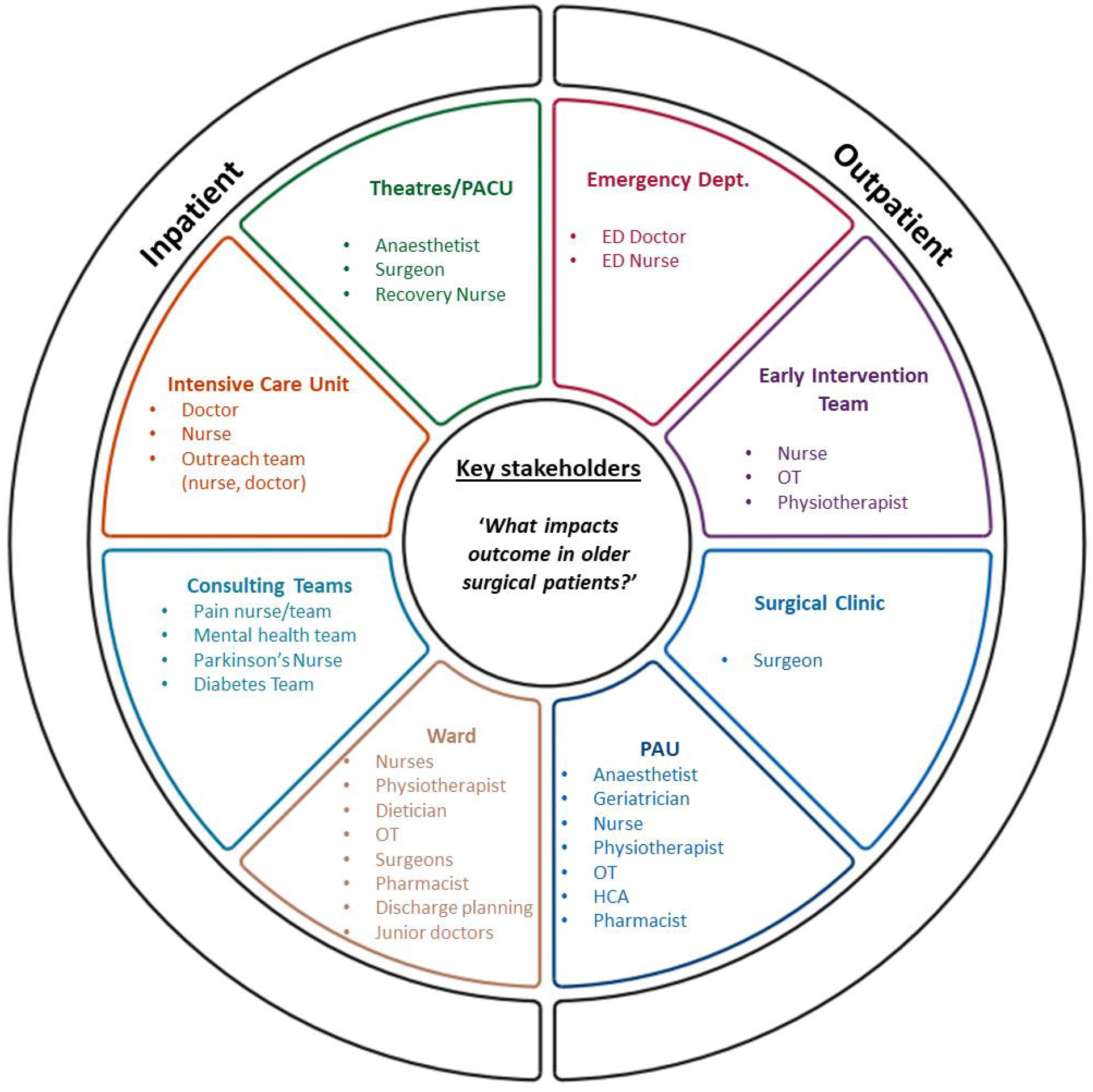
Stakeholder diagram: Stakeholder groups identified for representation on a Delphi panel to understand what defines a high-risk surgical patient and impacts on their outcome. All groups were identified by a collaborative process mapping exercise with a steering group of key perioperative professionals. Diagram template adapted from that available at www.iitoolkit.com, used with permission. *ED = Emergency Department, HCA = Health-care assistant, OT = Occupational therapist, PAU = Pre-assessment unit, PACU = Post-anaesthesia care unit*

### Stakeholders and Delphi participants

Invitations to participate were sent to 91 professionals, identified by leads in both trusts (63 from CUH 28 from WSH). These covered 23 broad professional groups as well as subspecialty expertise. Numbers of representatives are shown in **Supplementary Figure 3**. Participants were able to contribute views from more than one perspective (e.g. anaesthetist AND intensive care doctor). Response rates ranged from 51% (*n=46*) for the first round to 19% (*n = 17*) in the third round. Conduct of the Delphi was significantly impacted by the coronavirus pandemic - distribution of the second and third rounds was delayed by six months due to the first wave and the final round was completed as case pressures built prior to the second national UK lockdown in November 2020.

### Minimum dataset: variables from the literature

25 risk scores were identified from a 2013 systematic review [4]. A full list of scores and their component variables are demonstrated in **supplementary table 1** and **supplementary figure 3**. Any of the variables not suggested in round one were included by default for voting in rounds two and three.

### Delphi round one

From the literature, 168 variables (representing specific measurements or characteristics) were identified (**Supplementary figure 3**). From free-text responses from participants, 411 suggestions for variables were identified including 243 unique or refined definitions from those present in the literature. 80 of the 168 (48%) literature variables were not suggested by the panel in round one. Suggestions included

To provide context to questions in later rounds, all suggestions were grouped into a framework separating out suggestions pertaining to the *patient*, their *admission*, and the *organisation* caring for them. For clarity of questioning, suggestions were further organised into related groups (such as comorbidities), sub-groups (e.g. cardiovascular comorbidities), and then a granular level that represented specific concepts or definitions (e.g. ‘electrocardiogram changes’ and ‘left bundle branch block’). This structure is demonstrated in **Figure 4** and is available to view interactively in the **supplementary material**. Distribution of new suggestions across these domains of *patient, admission, and organisation* is shown in **Figure 5A**. None of the suggestions that appeared to pertain to health system organisation or performance (**Table 1**) featured in currently used risk scores (**Figure 5A)**.

**Figure 4:**
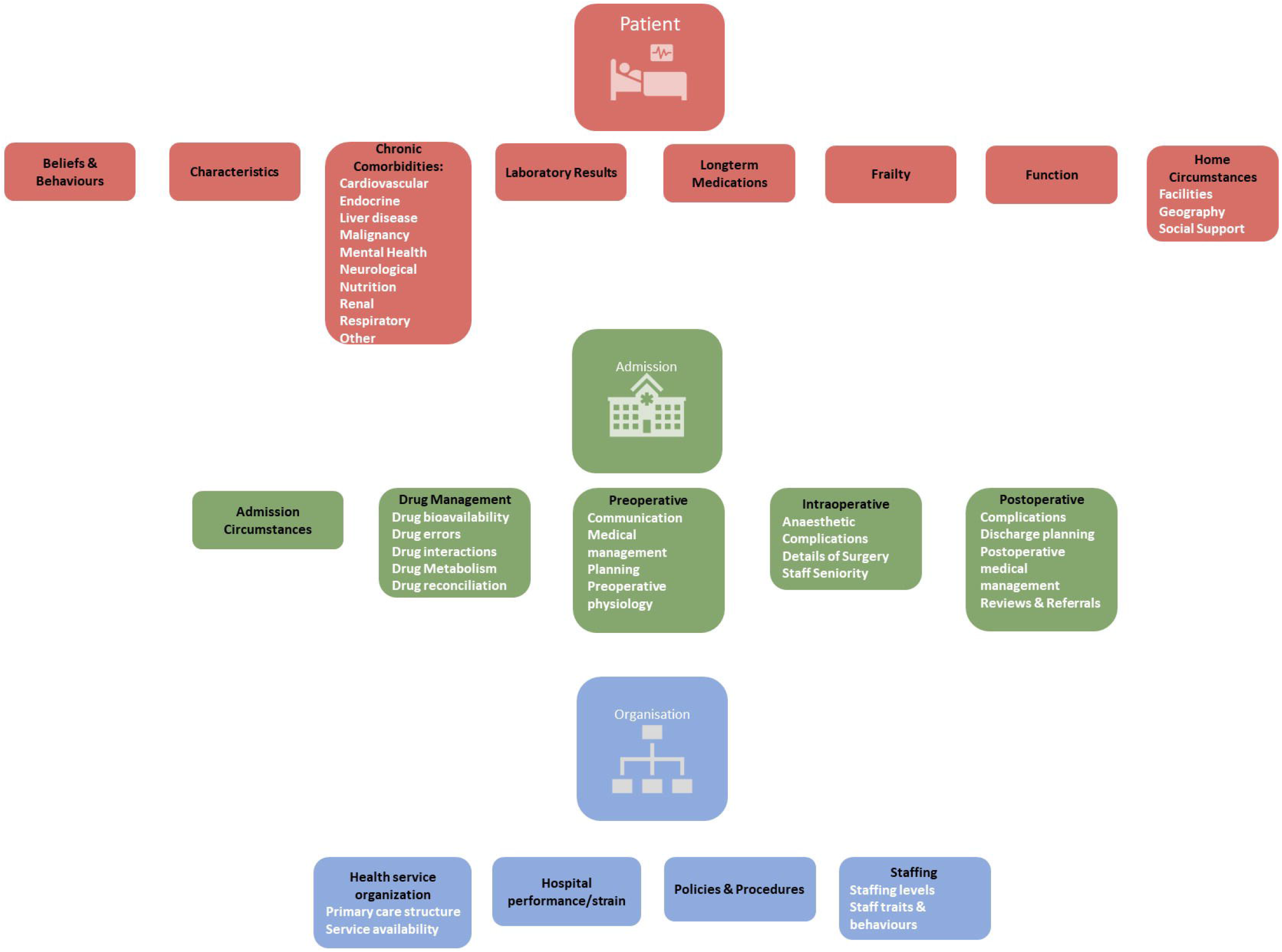
Delphi structure showing main risk factor groups identified through participant suggestions and the literature. The questionnaire enabled people to vote on three broad domains (Patient, Admission, Organisation) and relevant groups (black text in lozenges). Many of these groups had relevant subgroups (white text). Participants could also vote on individual date elements (variables or their definitions) within each of these. Full list of these are available in the supplementary material.

**Figure 5:**
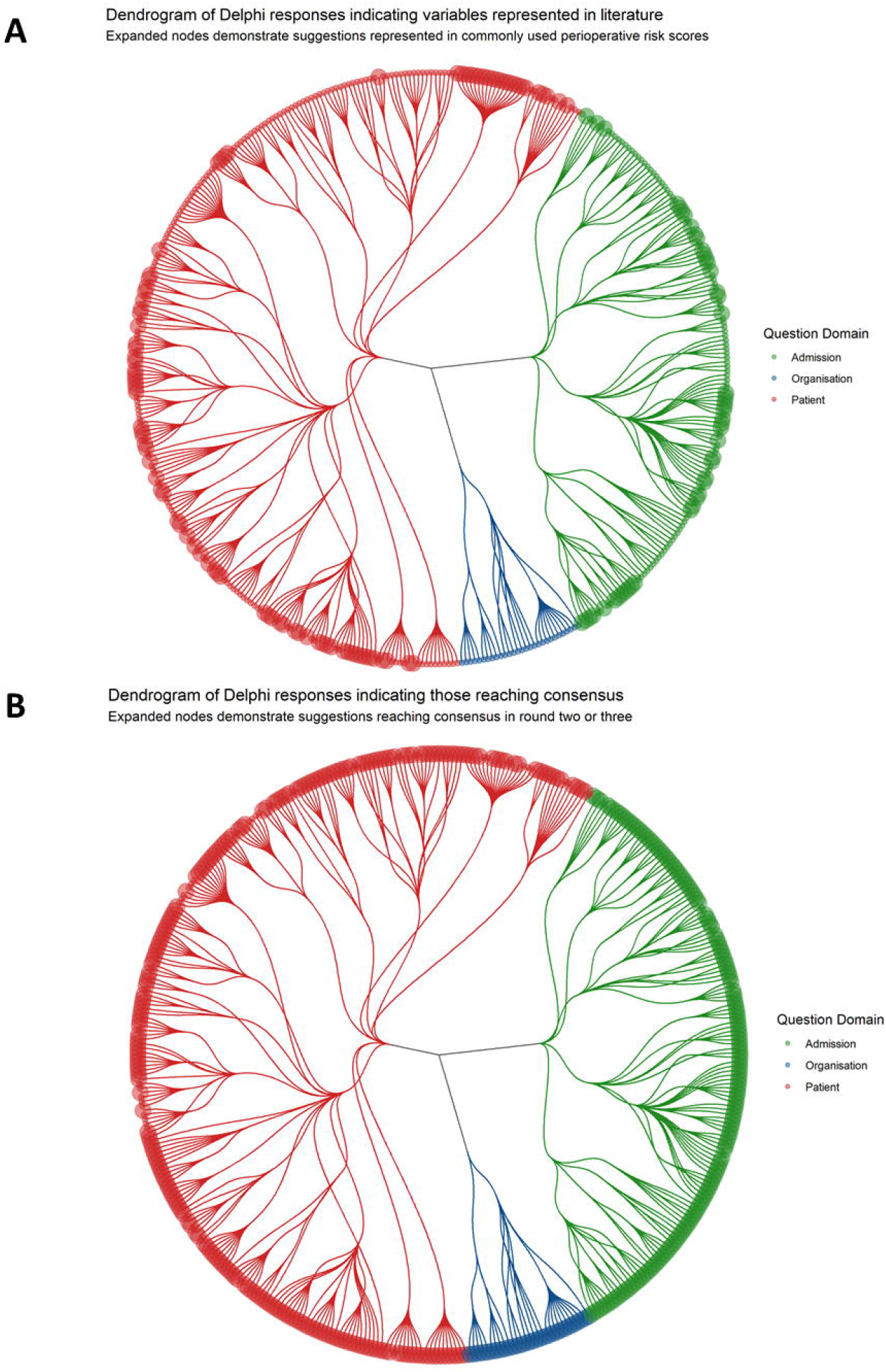
Representation of the Delphi questionnaires as a dendrogram, demonstrating hierarchical organisation of questions. Colours represent broad domains of statements being voted on by the panel; pertaining to a patient, circumstances of admission, or those pertaining to the organisation caring for them. Final questions that were voted on are represented by terminal leaves. Branch points represent question structure which can be viewed in the interactive html document within supplementary material. In panel **A**, expanded nodes represent those variables derived from a systematic review of commonly used perioperative risk scores. In panel **B**, expanded nodes represent those that reached criteria for consensus inclusion in a dataset seeking to understand drivers of perioperative risk in older people.

**Table 1.** Examples of ‘organisational’ factors suggested by Delphi participants as being important in the determination of outcome for older surgical patients. All of these suggestions were made by participants in the first Delphi round and reached consensus across round two or three.

| Suggested factor | Question group |
| --- | --- |
| Availability of services | Health Services Organisation |
| Number of patients cared for (*by each GP*) | Health Services Organisation |
| GP practice type | Health Services Organisation |
| Presence of a telephone advice service | Health Services Organisation |
| Tertiary admission/transfer | Health Services Organisation |
| Bed Occupancy Rates | Hospital Performance |
| Delayed Transfer of Care (DToC) Rates | Hospital Performance |
| ED Admission wait times | Hospital Performance |
| Intensive care occupancy | Hospital Performance |
| Number of outlying patients | Hospital Performance |
| Number of vacant posts | Hospital Performance |
| Overnight stay in ED | Hospital Performance |
| Respiratory Virus Rates | Hospital Performance |
| Season | Hospital Performance |
| Surgery cancellation rates | Hospital Performance |
| Care Pathway Documents | Policies and Procedures |
| Drug availability (e.g. anticoagulant reversal agents) | Policies and Procedures |
| Enhanced recovery programs | Policies and Procedures |
| Procedures for minimising conflicts between teams | Policies and Procedures |
| Procurement | Policies and Procedures |
| Empathy | Staffing |
| Enthusiasm and engagement | Staffing |
| Expertise in the care of older people | Staffing |
| Medical cover | Staffing |
| Nursing cover | Staffing |
| Occupational therapy cover | Staffing |
| Organisational culture | Staffing |
| Pharmacy cover | Staffing |
| Physiotherapy cover | Staffing |
| Staff traits, skills, behaviours | Staffing |
| Staffing levels | Staffing |
| Training levels | Staffing |

### Delphi rounds two and three

In round two suggestions were presented for voting with participants able to vote on all levels of the questionnaire hierarchy (**Figure 4**) including groups and subgroups. In total 409 suggestions including 17 groups, 34 subgroups, and 358 variables (of which 117 were operationalised definitions of a variable - such as ‘left bundle branch block’ representing an ‘important ECG change’) were voted on. Two suggestions from the risk scores (cut-off values for defining polypharmacy) were held until round three as free-text suggestions for this threshold were sought from panellists in round two. Full details of suggestions that were presented for voting across the final two rounds, including number of responses and consensus thresholds, are available as a **supplementary spreadsheet file**

At the conclusion of round two, 357 (87%) of statements reached consensus criteria for inclusion. Analysis of free text suggestions identified a further 108 suggested variables for voting in the third Delphi round.

The median score for measurability in round two was four [IQR:3-4]. Laboratory results were felt to be the most measurable (attracting a median score of five) with patient *beliefs and behaviours, health service organisation, hospital performance & strain*, and *policies & procedures* attracting median scores of three.

In the third Delphi round, 110 of 158 (70%) variables met the consensus criteria for inclusion, one variable (“*shortness of breath on strenuous exercise”*) met the criteria for exclusion. In total, across both rounds, 468 of 519 (90%) suggestions that were presented to the panel reached consensus decisions (**Figure 5B**). In round three median measurability was put at four [IQR:4-5] with only *beliefs & behaviours* and *health service organisation* attracting median scores of less than four. Across both latter rounds all of the suggestions encompassing health system organisation reached the definition for consensus.

### Comparison to literature variables

To assess whether suggested variables might have been previously examined but excluded from published risk scores due to a lack of statistical significance we identified the original papers generating each of the risk scores[19–32]. Twenty-five (7%) of the 351 unique suggestions from rounds one and two had been previously examined at an earlier phase in the development of at least one risk tool (details in **Supplementary spreadsheet**).

All novel suggestions were also compared against a list of process and structure indicators from a 2018 systematic review[18]. 106 of 351 (30%) novel suggestions could be mapped against one or more metrics identified in this paper. When new suggestions defining patient level characteristics (e.g., comorbidities) were excluded - 87 of 160 (54%) new suggestions relating to admission circumstances or organisational function were represented. Variable that did not appear included markers of system performance (e.g., *number of vacant posts, delayed transfer of care rates)* and staffing (e.g., *occupational therapy cover*) as well as examples of postoperative complications (e.g., *anastomotic breakdown*). For ease of reference these variables are shown in **supplementary table S2** but full details can be seen in the **supplementary spreadsheet file**.

## Discussion

This study demonstrates the views of multidisciplinary clinicians on factors felt to influence perioperative risk. We feel that both our findings and methods will be of interest to researchers within the perioperative field as well as in other disciplines seeking to rationally use EHRs for research and improvement. Our results highlighted factors that were not surgical or patient characteristics but instead related to in-hospital events, organisational structure, and hospital performance. Such suggestions were conspicuously absent from commonly used perioperative risk scores **(Figure 5A)** [4] but are compatible with work demonstrating inter-centre variation in outcome [7]. Our final list of consensus variables (available in supplementary) is likely to be of use to researchers in the field seeking to intelligently curate data from their own EHRs, the fact that the panel also voted on specific cut-points (e.g., ‘Clinical Frailty Scale of > 5’) may also be of use in those seeking to develop relevant triage points within guidelines.

Methodologically we hypothesised that process mapping may enable us to identify a panel of stakeholders whose expertise captured all facets of perioperative care and, that in doing so, we may gain novel insights. This approach is at least partially vindicated in that most factors voted on (351/519) were suggested by the panel. Future work is needed to validate the importance of these findings in understanding whether these in-hospital processes affect or predict patient outcome. An additional strand of research should explore how an understanding of system structure (captured using a process map) may aid researchers in appreciating how care processes may impact on broader data capture within the HER.

The importance of ‘non-patient’ level factors in prognostication has not been thoroughly explored, perhaps reflecting the challenges in recording such concepts electronically. When considering quantitative data from EHRs, improved risk assessment can be seen by incorporating implicit information such as the timing of blood samples, as well as their resulting value [34]. Here, this timing information is presumably recognising a potentially otherwise undocumented recognition of a clinically unwell patient by a diligent clinician taking, and interpreting, blood samples out of hours. The results of this study suggest that variables not captured in a hospital EHR, such as detailed aspects of social circumstances, may be important. A machine learning model containing only ‘sociomarkers’ (derived from place of residence) has yielded comparable performance in the prediction of childhood asthma exacerbation compared to the use of patient-level data [35]. However, conflicting findings have been shown in other settings. Although qualitative data derived from patient interviews has suggested that social support and psychological state are influences on individual readmissions with heart failure[36], small studies looking to incorporate questionnaire derived markers have failed to demonstrate improved prognostication [37,38] despite the apparent use of ‘unstructured’ free-text data in another study [39]. Perioperatively it is well known that clinician judgement improves the performance of commonly utilised risk tools such as SORT (Surgical Outcome Risk Tool) [40]. What is unclear is what additional factors this clinician judgement encapsulates and whether it may reflect an appreciation or implicit judgement of some of the factors identified in this study.

A prerequisite for validating these suggestions in future work is identifying whether valid electronic surrogates exist. One approach to try and address this may be to broaden the definition as to what we view as constituting ‘healthcare data’. The Institute of Medicine suggests that relevant ancillary data sources may include human resources records and patient complaints [41]. This could conceivably aid with the incorporation of factors such as staffing levels with complaints data, if adequately coded using tools such as the Healthcare Complaints Analysis Tool (HCAT), identifying poor staff-patient relationships and communication [42].

This study took the unique step of drawing on techniques from systems engineering to structure a Delphi survey of professionals. These methods will be of direct relevance to those involved in informatics, research, or quality improvement both within and beyond perioperative practice. The methods and results of this work also reflects the relevance of systems thinking in healthcare. A systems approach is a way of addressing problems holistically, aware of the interaction between elements and subsequent unexpected behaviour[11,43]. Suggestions from the panel related to the circumstances of admission, hospital performance, and external pressures. A conceptual framework is thus that the ultimate outcome of any patient stay depends on the relative interaction between factors at different levels. Conceptually this hierarchical agreement is appealing with the importance of external factors, such as coronavirus pressures, a key part of current daily practice.

Beyond our unique methodological approach we feel that a further point of originality within our work is that the Delphi panel were specifically asked to suggest factors that could result in a loss of independence on discharge, a key concern of patients undergoing surgery [44]. Despite this it is not widely considered in perioperative risk stratification[4]. An additional strength of our results is the multidisciplinary nature of the Delphi panel with a plurality of perspectives offering detailed insight into potentially important factors across the perioperative pathway, from drug management to home circumstances and medical management.

We do however acknowledge the fall off in response rate across rounds (51% in round one to 19% in round three). This arose due to the significant pressures of the coronavirus pandemic and that many stakeholder groups on our panel were undertaking additional clinical duties and redeployment to intensive care. However, we would counter this by highlighting the breadth of specialties surveyed, and that the response rate in round two was still 38% and that a majority of consensus factors (357/490) were identified in this round. It is possible that the low response rate in round three skewed the remaining factors towards reaching consensus, but this should make our resultant list of variables especially sensitive (albeit potentially less specific). The high rates of consensus could also reflect true strength of feeling but may have arisen due to issues with questionnaire length, panel composition, and response rates.

## Conclusion

This study demonstrates the feasibility of using systems engineering tools to identify and engage clinicians in identifying factors felt to impact on patient relevant outcomes after surgery. The results themselves highlight that these professionals identify non-patient level factors as modulators of perioperative risk. Further work is needed to develop electronic surrogates for these suggestions and validate their significance in real datasets so that their relevance as drivers or markers of unwarranted variation can be assessed.

## Supporting information

which can be viewed in the html document

## Data Availability

In Supplementary material

## Acknowledgements

This research was funded, in whole or in part, by the Wellcome Trust, Grant number: 220542/Z/20/Z (to DJS). A CC BY or equivalent licence is applied to the Author Accepted Manuscript (AAM) arising from this submission, in accordance with the grant’s open access conditions.

PJC and NL both contributed equally to this work and should be considered joint senior author

This research was supported by the NIHR Cambridge Biomedical Centre (BRC 1215 20014). The views and opinions expressed by authors in this publication are those of the authors and do not necessarily reflect those of the NHS, the NIHR, or the department of health.

## Data Sharing Statement

Summary results for each round of our study are available in the supplementary material.

## Competing Interests Statement

No competing interests declared

## Notes

### Competing Interest Statement

The authors have declared no competing interest.

### Author Declarations

London and Surrey Borders NHS Research Ethics Committee (reference 19/L0/1648)

