## Supplementary material for "Can process mapping and a multi-site Delphi of perioperative professionals inform our understanding of system-wide factors that may impact operative risk?"

This document contains supplementary figures, **further material is available as two separately available files.**

1. A **.xlsx format** spreadsheet showing all suggestion voted on across the three rounds of the Delphi
2. A **html format** document with an interactive collapsible view of the hierarchy of questions used in the second and third delphi rounds. Details on how to browse this can be found in section 6 of this document

### Contents of this document

1. **Annotated process maps demonstrating the care of elective (a) and emergency (b) vascular surgical patients**
2. **List of risk scores used to provide a ‘minimum’ dataset for voting on in Delphi round two**
3. **Component variables from these risk scores**
4. **Hierarchy of question groupings used in Delphi rounds two and three**
5. **‘Novel’ variables from the Delphi**
6. **Using the interactive html file**

**Supplementary Figure 1: Process map of elective vascular care used to identify perioperative stakeholders.**
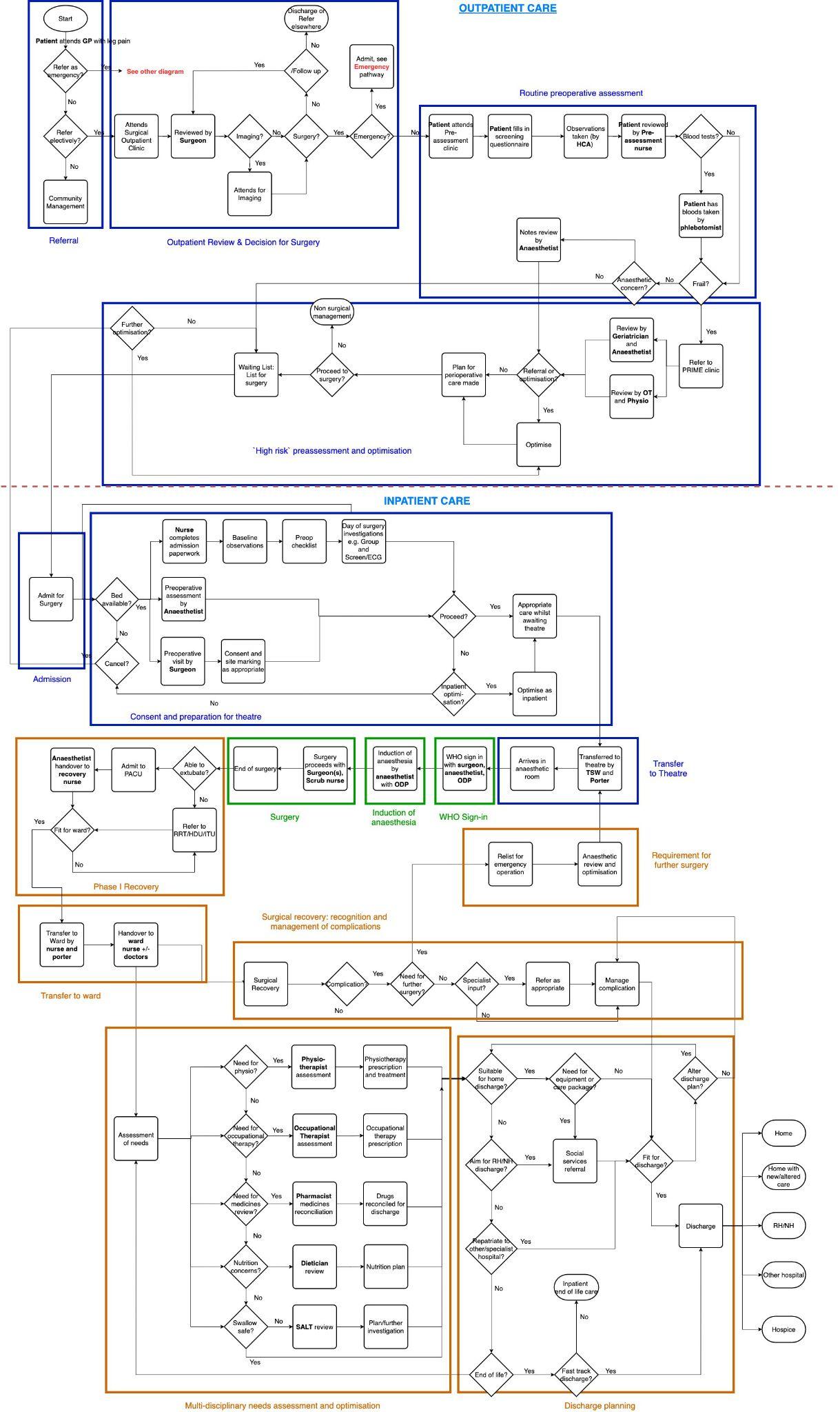

**Supplementary Figure 2: Process map of emergency vascular care used to identify perioperative stakeholders. \**
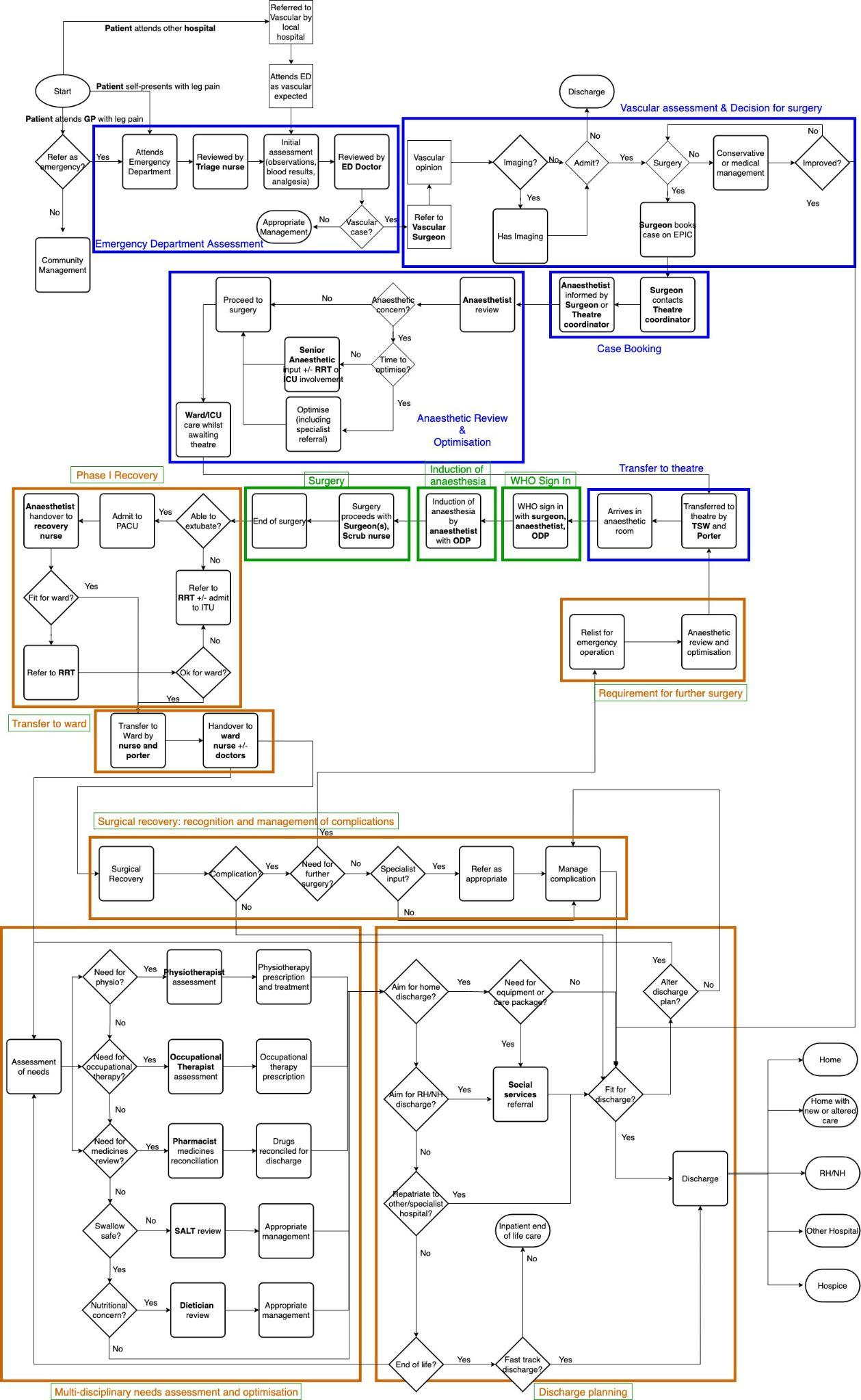

(In both Figure S1 and S2, coloured boxes correspond to abridged care processes shown in Figure 1 in main text)

**Risk scores used in formation of a ‘minimum’ dataset for voting in rounds two and three**

**ASA score Nutritional risk index**

**BHOM Otago Surgical outcome score**

**Charlson Comorbidity Index Perioperative Mortality Risk Score**

**APACHE POSSUM**

**Delaware P-POSSUM**

**Detsky index RCRI**

**E-PASS SAPS II**

**Edmonton Frail scale Subjective Global assessment**

**ICISS Surgical Apgar**

**IRIS Surgical mortality score**

**Maastricht Nutritional Assessment Surgical Risk Scale**

**Mini nutritional assessment Surgical Risk Score (Donati)**

**MPM III**

**Supplementary Table 1:** List of risk scores identified as having been validated in heterogeneous surgical cohorts in a 2013 systematic review[[1]](https://paperpile.com/c/1RqjnS/Fsg4). These scores were deconstructed (see **Supplementary Figure 3**) to provide the minimum components of any dataset resulting from the Delphi. *ASA = American Society of Anesthesiologists,*

*BHOM – Biochemistry & Haematology Outcome Model, APACHE = Acute physiology & chronic*

*health evaluation, DELAWARE = Dense Laboratory Whole blood applied Risk Estimation, E-PASS = Estimation*

*of physiological ability and surgical stress, ICISS = ICD injury severity score, IRIS = Identification of risk in*

*surgical patients score, MPM III = Mortality prediction model (III), POSSUM = Physiological and Operative*

*Severity Score for the enUmeration of Mortality and morbidity, P-POSSUM = Portsmouth modification of*

*POSSUM, RCRI = Revised cardiac risk index, SAPS II = Simplified acute physiology score II*

1. Moonesinghe SR, Mythen MG, Das P, Rowan KM, Grocott MP. Risk stratification tools for predicting morbidity and mortality in adult patients undergoing major surgery: qualitative systematic review. *Anesthesiology* 2013; **119**: 959–81.

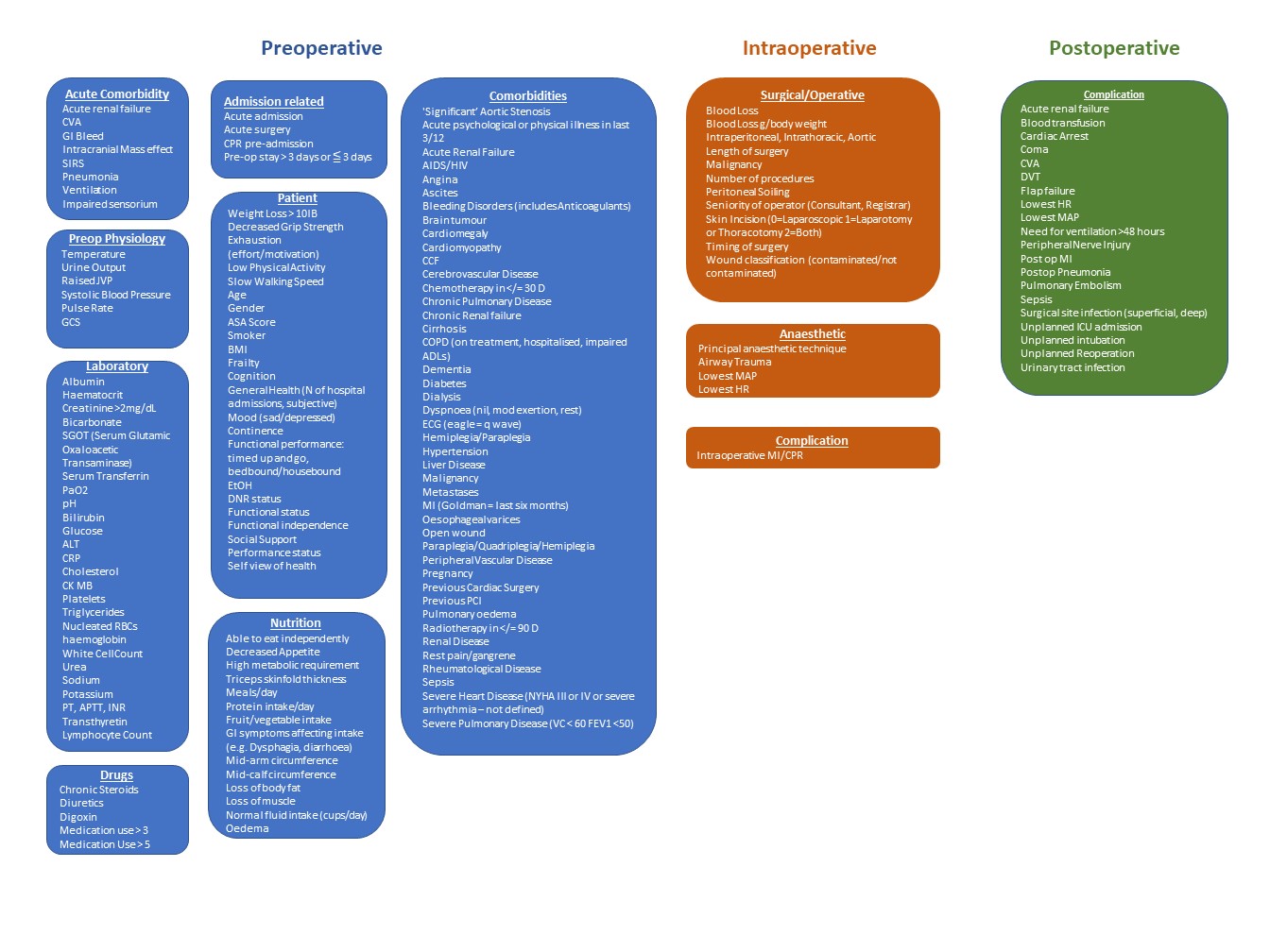

**Supplementary Figure 3: Components of validated perioperative scores introduced into Delphi at round two.** These variables were identified by deconstructing the risk scores shown in Supplementary Table 1. Groupings are artefactual for clarity and to delineate key temporal aspects of the use of the identified risk scores. Each of these variables were included in Delphi round two, regardless of whether they were present in free-text suggestions from the panel in round one.

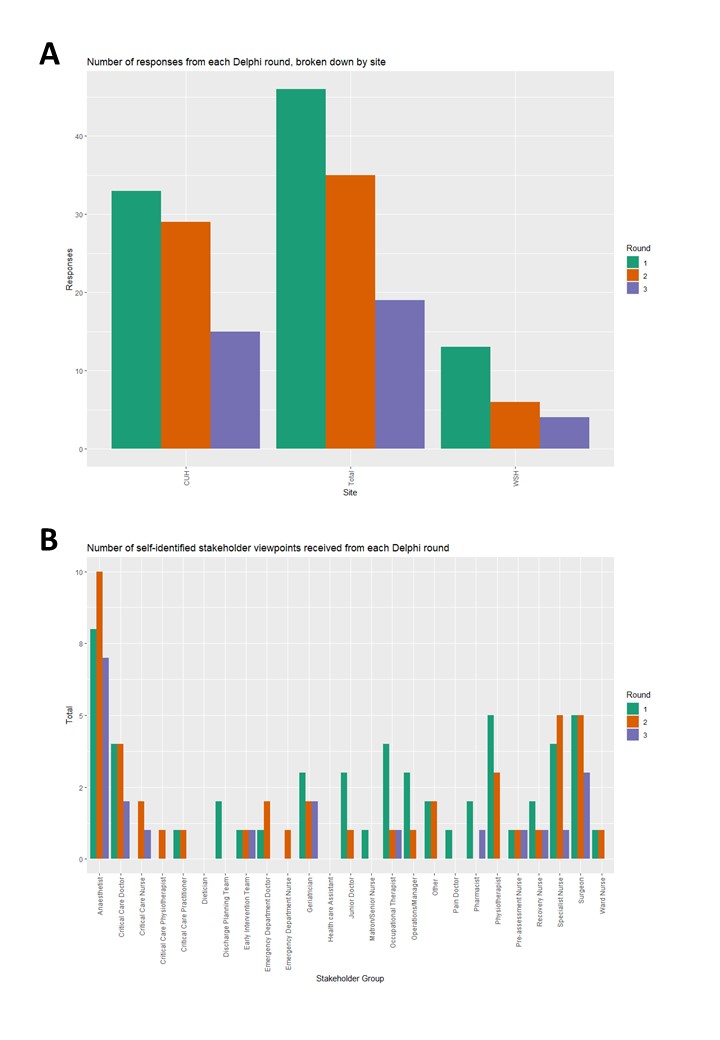

**Figure 3: Response rate and respondent profile** **by round for a Delphi exercise to gain a consensus view on factors impacting on outcome in older surgical patients.** *CUH = Cambridge University Hospitals (a tertiary care centre), WSH = West Suffolk Hospital (a secondary care centre)*

**Variables suggested by the Delphi panel that did not feature in the development of perioperative risk scores or in a systematic review of structural and process quality indicators.**

| **VARIABLE** | **DOMAIN** | **GROUP** |
| --- | --- | --- |
| **Opioid deprescribing pre-discharge** | Admission | Analgesic Plan |
| **Opioid dose** | Admission | Analgesic Plan |
| **Chest injuries** | Admission | Circumstances |
| **Delayed presentation** | Admission | Circumstances |
| **Disruption to a patient's routine** | Admission | Circumstances |
| **Bed Occupancy Rates** | Organisation | Hospital Performance |
| **Subdural haematoma** | Admission | Circumstances |
| **Trauma** | Admission | Circumstances |
| **Indication for surgery (pathology)** | Admission | Details of Surgery |
| **Orthopaedic v Non-Orthopaedic** | Admission | Details of Surgery |
| **Delayed Transfer of Care (DToC) Rates** | Organisation | Hospital Performance |
| **Delayed Transfer of Care** | Admission | Discharge Planning |
| **ED Admission wait times** | Organisation | Hospital Performance |
| **Empathy** | Organisation | Staffing |
| **Enthusiasm and engagement** | Organisation | Staffing |
| **Health Service Organisation** | Organisation | Health Services Organisation |
| **Hospital Performance & Strain** | Organisation | Hospital Performance |
| **Intensive care occupancy** | Organisation | Hospital Performance |
| **Details of Surgery** | Admission | Intraoperative |
| **Intraoperative CO2 Control** | Admission | Intraoperative |
| **Intraoperative Complications** | Admission | Intraoperative |
| **Medical cover** | Organisation | Staffing |
| **Intraoperative Physiological Control** | Admission | Intraoperative |
| **ODP Seniority** | Admission | Intraoperative |
| **Operator Fatigue** | Admission | Intraoperative |
| **Scrub nurse Seniority** | Admission | Intraoperative |
| **Number of outlying patients** | Organisation | Hospital Performance |
| **Number of patients cared for** | Organisation | Health Services Organisation |
| **Number of vacant posts** | Organisation | Hospital Performance |
| **Nursing cover** | Organisation | Staffing |
| **Occupational therapy cover** | Organisation | Staffing |
| **Escalation plan made** | Admission | Postoperative |
| **Organisational culture** | Organisation | Staffing |
| **Overnight stay in ED** | Organisation | Hospital Performance |
| **Number of attachments** | Admission | Postoperative |
| **Pharmacy cover** | Organisation | Staffing |
| **Physiotherapy cover** | Organisation | Staffing |
| **Referrals & Reviews** | Admission | Postoperative |
| **Time to occupational therapist review** | Admission | Postoperative |
| **Time to pharmacist review (e.g. for medicines reconcilliation)** | Admission | Postoperative |
| **Anastomotic Breakdown** | Admission | Postoperative Complications |
| **Practice type** | Organisation | Health Services Organisation |
| **New diagnosis of chronic conditions (e.g. Dementia)** | Admission | Postoperative Complications |
| **Poor Wound Healing** | Admission | Postoperative Complications |
| **Postoperative Dysrhythmia** | Admission | Postoperative Complications |
| **Postoperative Pain** | Admission | Postoperative Complications |
| **Postoperative Pneumonia** | Admission | Postoperative Complications |
| **Postoperative sepsis** | Admission | Postoperative Complications |
| **DNR status & Escalation plan** | Admission | Preoperative |
| **How long patients expect to stay in hospital** | Admission | Preoperative |
| **Primary Care Structure** | Organisation | Health Services Organisation |
| **Procurement** | Organisation | Policies and Procedures |
| **Prehabilitation program** | Admission | Preoperative |
| **Respiratory Virus Rates** | Organisation | Hospital Performance |
| **Season** | Organisation | Hospital Performance |
| **Therapy plans** | Admission | Preoperative |
| **Staffing** | Organisation | Staffing |
| **Staffing levels** | Organisation | Staffing |
| **Able to follow commands preoperatively** | Admission | Preoperative Physiology |
| **Tertiary admission/transfer** | Organisation | Health Services Organisation |
| **Preoperative CVS impairment** | Admission | Preoperative Physiology |
| **Preoperative Delirium** | Admission | Preoperative Physiology |
| **Preoperative Physiology** | Admission | Preoperative Physiology |
| **Preoperative Renal impairment** | Admission | Preoperative Physiology |
| **Preoperative Respiratory Failure** | Admission | Preoperative Physiology |
| **Ward moves/handovers** | Admission | Postoperative |

*Supplementary Table 2:* List of variables that emerged during the Delphi that did not feature in the development of perioperative risk scores or in a systematic review of quality and structural process indicators. For references see text. Blue highlighting demonstrates factors related to the organisation of the healthcare organisation, Green highlights factors pertaining to the individual admission in question.

**Using the Interactive Html File**

An interactive html file **(DelphiQViewer.html)** is available as a separate supplementary file. By clicking on the interactive diagram you will be able to view the hierarchy used in presenting all options to the panelists for voting. The lowest level (or ‘leaf’) is the most granular level of suggestion that the panelists voted on.

**Click** on each node to expand lower levels, white nodes demonstrate the lowest level of each branch. As new branches expand **click and drag** on the surrounding area to re-center the diagram as you go. Zooming in and out can be achieved using the **scroll wheel** of your mouse.
